# Predicting the impact of non-pharmaceutical interventions against COVID-19 on *Mycoplasma pneumoniae* in the United States

**DOI:** 10.1101/2024.08.19.24312254

**Authors:** Sang Woo Park, Brooklyn Noble, Emily Howerton, Bjarke F Nielsen, Sarah S Jiudice, Lilliam Ambroggio, Samuel Dominguez, Kevin Messacar, Bryan T Grenfell

## Abstract

The introduction of non-pharmaceutical interventions (NPIs) against COVID-19 disrupted circulation of many respiratory pathogens and eventually caused large, delayed outbreaks, owing to the build up of the susceptible pool during the intervention period. In contrast to other common respiratory pathogens that re-emerged soon after the NPIs were lifted, longer delays (*>* 3 years) in the outbreaks of *Mycoplasma pneumoniae* (Mp), a bacterium commonly responsible for respiratory infections and pneumonia, have been reported in Europe and Asia. As Mp cases are continuing to increase in the US, predicting the size of an imminent outbreak is timely for public health agencies and decision makers. Here, we use simple mathematical models to provide robust predictions about a large upcoming Mp outbreak in the US. Our model further illustrates that NPIs and waning immunity are important factors in driving long delays in epidemic resurgence.

## 1 Introduction

*Mycoplasma pneumoniae* (Mp) is the most commonly detected bacterium for lower tract respiratory infections in children and adults (Waites and Talkington, 2004; Jain et al., 2015,?; Bajantri et al., 2018). Mp pneumonia can affect a patient for a long period due to its long incubation period and long prodromal duration of symptoms, especially among children and high risk individuals. For instance, outbreaks in schools have resulted in increased hospitalization, ventilator-associated pneumonia, severe extrapulmonary disease (e.g. Stevens-Johnson syndrome) and have been shown to last for several months (Walter et al., 2008; Olson et al., 2015). Increasing levels of macrolide-resistant Mp strains further highlight the importance of appropriate diagnosis and treatment (Pereyre et al., 2016).

Multiannual cycles in Mp outbreak patterns have been observed in many countries, with large outbreaks occurring every 3–7 years (Kim et al., 2009; Brown et al., 2016). Several potential mechanisms have been proposed to explain the observed epidemic cycles, including waning immunity (Omori et al., 2015) and strain dynamics (Kenri et al., 2008; Zhang et al., 2019). More parsimoniously, multiennial epidemic dynamics can also arise from simple, seasonally-forced immunizing epidemic dynamics, especially given low transmission rates (Earn et al., 2000; Keeling et al., 2001).

Along with other endemic viruses and bacteria, the circulation of Mp was disrupted in 2020 due to non-pharmaceutical interventions (NPIs) that were introduced to prevent the transmission of SARS-CoV-2 (Boyanton Jr et al., 2024). The disruption of the expected epidemiological curve adds challenges to predicting future Mp outbreaks. In contrast to other common respiratory pathogens that re-emerged within a year after NPIs were lifted, the re-emergence of Mp outbreaks has been delayed for more than 3 years in Europe and Asia (Sauteur et al., 2024). A long delay in epidemic resurgence is particularly alarming because it can allow for a build up of a large susceptible population, increasing the risk of a large outbreak as the infection resurges (Baker et al., 2020).

As Mp infections are beginning to increase rapidly in the US (Figure 1A), predicting the size of an impending outbreak is critical for public health agencies and decision makers. Here, we present a modeling analysis of Mp outbreaks in the US using data from 2015 onward and predictions for potential upcoming outbreaks and subsequent epidemic dynamics.

**Figure 1:**
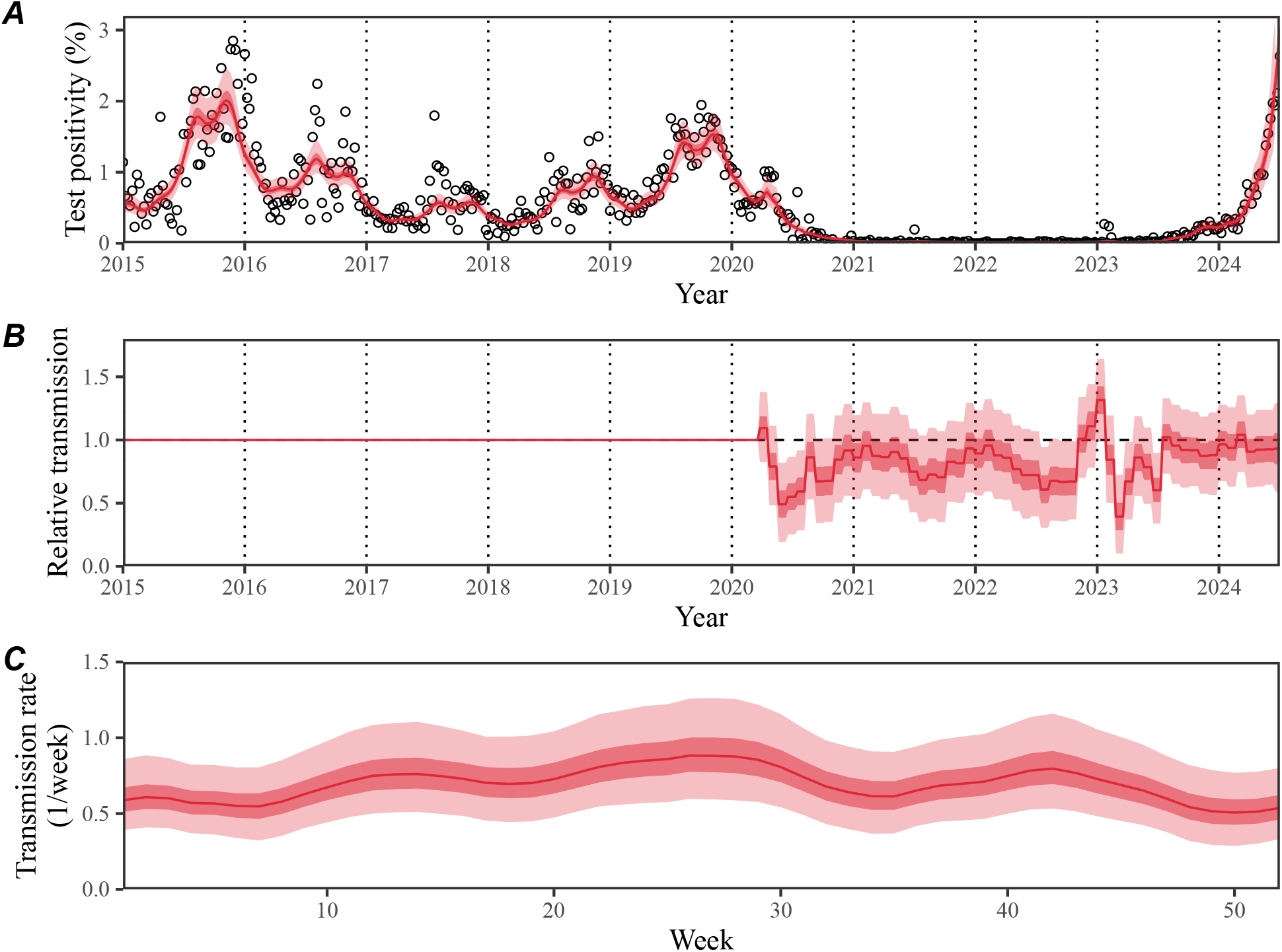
Summary of model fits to Mycoplasma pneumoniae positivity in the US, 2015–2024. (A) Comparisons of observed (points) and fitted (line) trajectories of weekly test positivity for Mp infections. (B) Estimated non-periodic, time-varying transmission term, representing relative transmission *δ* following the introduction of NPIs; for example, 0.5 corresponds to a 50% reduction in transmission. (C) Estimated periodic transmission term *β*_seas_(*t*), representing seasonal transmission rate. Red lines and shaded regions represent the estimated posterior median and corresponding 95% and 50% credible intervals.

## 2 Methods

### 2.1 Data

We analyzed over 2.54 million BIOFIRE@ Respiratory (RP1.7, RP2, and RP2.1) Panel (bioMérieux, Salt Lake City, Utah) multiplex PCR test results for positive detections of Mp (Poritz et al., 2011; Leber et al., 2018; Creager et al., 2020). These deidentified patient test results were captured from 127 US facilities from January 1, 2015 to June 29, 2024 using BIOFIRE@ Syndromic Trends, a cloud-based pathogen surveillance network for BIOFIRE@ instruments (Meyers et al., 2018).

### 2.2 Mathematical modeling of Mp outbreaks

To estimate the epidemic dynamics of Mp infections in the US, we fitted a seasonally-forced, deterministic Susceptible–Infectious–Recovered-Susceptible (SIRS) model (Pons-Salort and Grassly, 2018) to Mp time series data. The SIRS model considers three compartments, each representing the current infection status of an individual: Susceptible, Infected, and Recovered. We assumed a homogeneously mixed population, where infected individuals (I) transmit infection to susceptible individuals (S) at rate *β*(*t*) and recover at rate *γ* = 1*/*3 weeks (Omori et al., 2015). Recovered individuals were assumed to return to a susceptible state at rate *ν*, which we estimated by fitting the model to data. Birth and death rates *μ* = 1*/*80 years were assumed to be equal to fix the population size. In order to capture epidemic dynamics before and after NPIs were introduced, we extended the SIRS model by decomposing the transmission rate *β*(*t*) into two separate terms: (1) a periodic term with a period of 1 year *β*_seas_(*t*) that captures seasonal transmission, reflecting epidemic peaks in summer and fall (Figure 1A) and (2) a non-periodic, time-varying term that captures changes in contact patterns after the introduction of COVID-19 NPIs in March, 2020 *δ*(*t*):

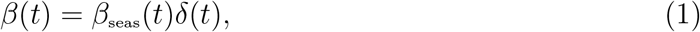

where *δ <* 1 corresponds to reduction in transmission due to NPI effects. For the period term, we estimated a separate transmission rate for each of the 52 weeks in a year, while constraining the smoothness using random-walk priors. For the non-period term, we allowed *δ*(*t*) to vary every 4 weeks from March 2020 and assumed *δ*(*t*) = 1 prior to March 2020; we also assumed *δ*(*t*) = 1 for longer-term predictions (for *t* outside the data range). Both terms were directly estimated by fitting the model to data using Bayesian framework (Supplementary materials). Parameter estimation was performed in a Bayesian framework using the Hamiltonian Monte Carlo algorithm through the R package rstan (Carpenter et al., 2017; Stan Development Team, 2024). The resulting posterior distribution was used to predict future Mp outbreaks by projecting the model forward. As a sensitivity analysis, we also performed the analysis using the SIR model, which assumes that infection provides life-long immunity.

### 2.3 Evaluating the impact of NPIs and waning immunity on the delays in re-emergence of Mp outbreak

To evaluate the potential impact of NPIs on long delays on Mp outbreaks, we performed sensitivity analyses on how reduction in transmission affect the timing of Mp outbreak resurgence. Specifically, we separately varied the strength and duration of NPIs by modulating the estimated NPI effects *δ*(*t*). First, the strength of NPIs were modified by taking fold changes in transmission *δ* − 1 and scaling it by a factor of *θ* such that the resulting transmission rate corresponds to

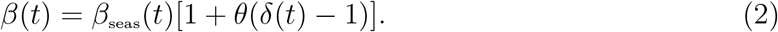

Second, the duration of NPIs were modified by assuming *δ* = 1 after the end date *t*_end_:

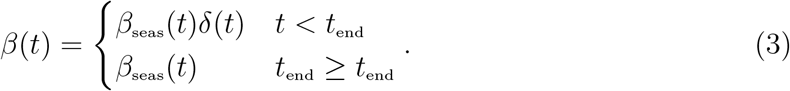

We varied *t*_end_ between 2020–2024. We note that we evaluate the effects of strength and duration of NPIs independently and do not explore their joint effects. Finally, we varied the duration of immunity 1*/ν* between 3–12 years to assess how waning immunity contributes to the timing of Mp outbreak resurgence.

## 3 Results

### Mathematical modeling of past Mp epidemics

The model reproduced the observed epidemic dynamics for Mp infections, including the seasonal and longer-term (≈ 5 year) epidemic cycles before NPIs were introduced and the delayed resurgence of the epidemic (Figure 1A). To capture the response of Mp to the pandemic, the model required a strong reduction in transmission (*δ*(*t*) *<* 1) in 2020 (Figure 1B). The model further estimated sustained reduction in transmission for 2021–2023 (Figure 1C). While we did not find a statistical correlation between the estimated changes in transmission and mean changes in Google mobility measures (*r* = −0.06; 95% CI: -0.22, 0.11), the strength of correlation increased with lags, with strongest correlation occurring at a 5-week lag (*r* = 0.25; 95% CI: 0.08–0.40; Supplementary Figure S1). The model demonstrated biannual patterns in seasonal transmission, peaking around weeks 26 and 42 (Figure 1C). The model also estimated the mean duration of immunity to be 9.3 years (95% CI: 4.4 years–16.4 years). The SIR model gave nearly identical estimates for the NPI effects but underestimated the peak of the 2015 outbreak, providing support for the SIRS model (Supplementary Figure S2).

### Prediction of future Mp outbreaks

In line with other acute, partially immunizing respiratory pathogens (Baker et al., 2020), the model predicted a large build-up of the susceptible pool during the NPI period (Figure 2A). This build up of susceptible individuals could lead to a large Mp outbreak, which appears to have begun (Figure 1A); the model forecasted that the epidemic would peak around the second half of 2024 and start to decline towards the end of 2024 (Figure 2B). The estimated peak positivity will be 7.5% (95% CI: 5.6–9.3%), which is 2–3 times larger than the observed peak positivity of 2.8% at the end of 2015. The model further predicted that the outbreak beginning in 2024 would cause a large depletion of the susceptible pool (Figure 2A), which would lead to another delayed outbreak after several years (Figure 2B).

**Figure 2:**
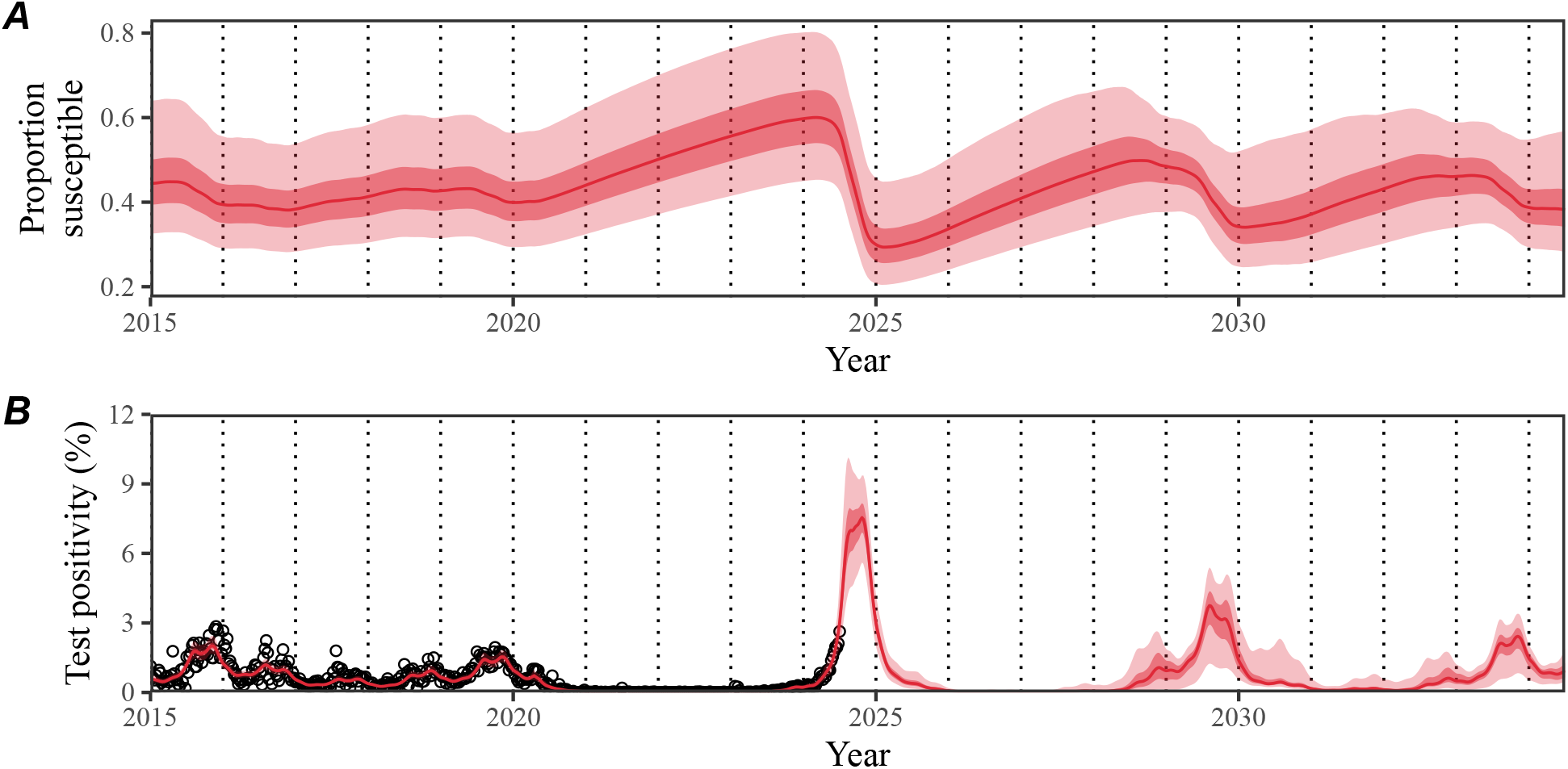
Predictions of future Mycoplasma pneumoniae outbreaks. (A) Predicted changes in the proportion of susceptible individuals. (B) Predicted changes in weekly test positivity for Mp infections. Red lines and shaded regions represent the estimated posterior median and corresponding 95% and 50% credible intervals.

### Impact of NPIs and waning immunity on the timing of Mp outbreak resurgence

The model predicted that a strong reduction in transmission due to NPIs is critical to explaining long delays in the Mp outbreak resurgence. A smaller reduction in transmission (Figure 3A) would have led to more persistent epidemics as well as earlier resurgence of Mp outbreaks (Figure 3B). Interestingly, the model still predicted a large outbreak in 2024 even when we assumed that NPI effects were 25% weaker than what we originally estimated, demonstrating the risk of a susceptible build up. Similarly, assuming shorter duration of NPIs (Figure 3C) led to earlier resurgence (Figure 3D). However, even if transmission rates were to return to pre-pandemic values by the end of 2020, the model predicted that the resurgence would not be observed until 2022 (Figure 3D). Finally, we found that the duration of immunity was also a key factor in determining the delays in Mp outbreak resurgences. Faster waning of immunity would have caused a faster build up of the susceptible pool, leading to an earlier Mp outbreak.

**Figure 3:**
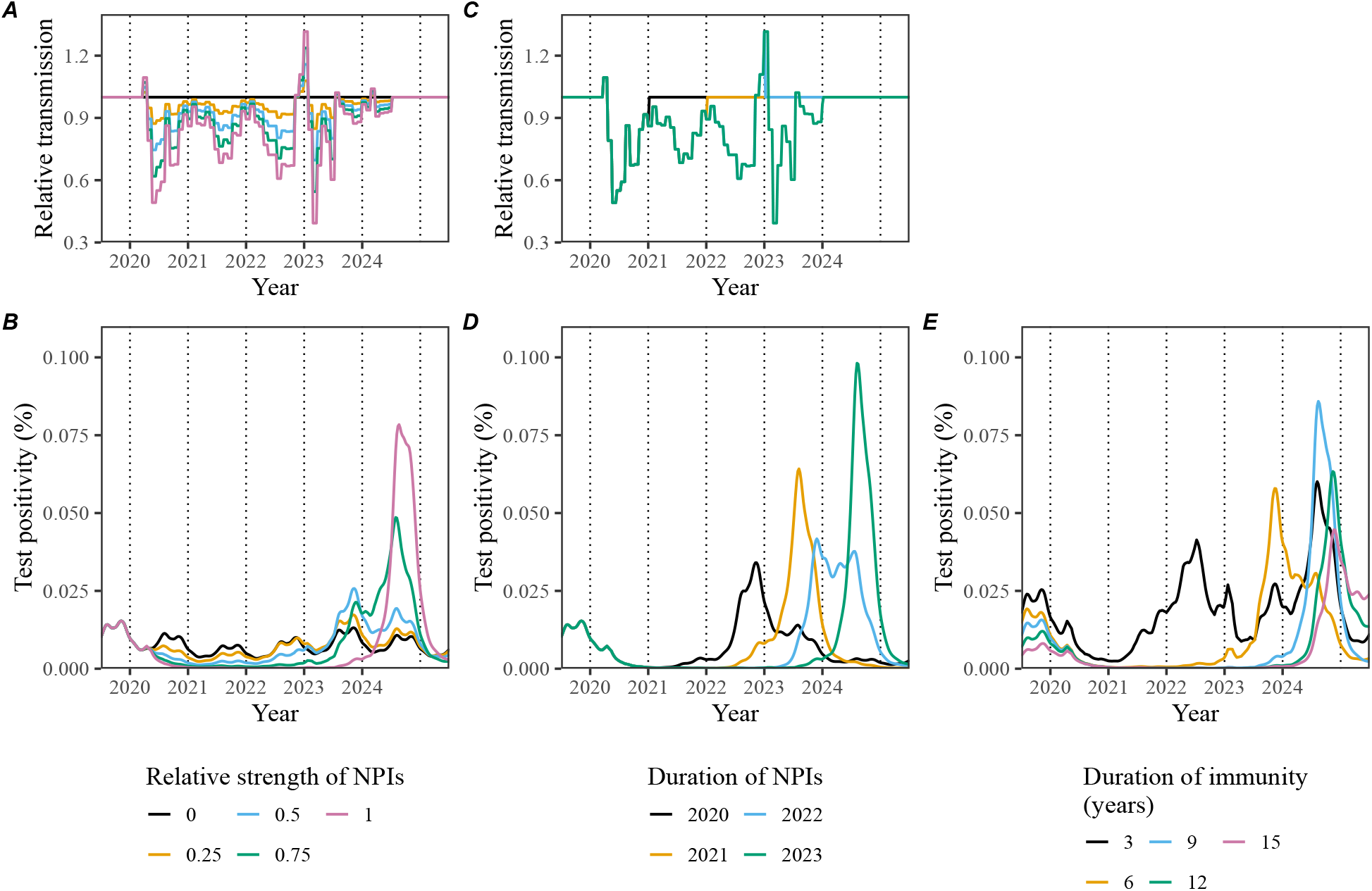
Impact of strength and duration of NPIs on the timing of Mp outbreak resurgence. (A, C) Assumed values for the relative transmission term *δ* following the introduction of NPIs by varying the strength (A) and duration (C) of NPIs. (B, D) Resulting epidemic dynamics across different assumptions about NPIs. (E) Resulting epidemic dynamics across different assumptions about the duration of immunity.

## 4 Discussion

In this study, we investigated the impact of NPIs on future Mp outbreaks in the US. By fitting a mathematical model to syndromic surveillance data, we predicted that a large Mp outbreak is imminent with a peak expected before the end of 2024. The upcoming outbreak is expected to be much larger than past outbreaks and may last until the end of 2025; note, however, that there is currently substantial uncertainty associated with longer-term dynamics, including the duration of 2024–2025 outbreak and the timing of the subsequent outbreaks. We can already observe what seems to be the beginning of this outbreak with an increasing rate of Mp detections throughout May and June 2024. Nonetheless, this prediction should alert clinicians and health care systems to be prepared for an increase in cases of pneumonia and potentially more rare presentations of Mp, such as reactive infectious mucocutaneous eruption (RIME) and encephalitis.

Our analysis highlighted the importance of strong reduction in transmission in explaining the delayed resurgence of Mp outbreaks. Specifically, we estimated ≈ 50% reduction in transmission in 2020, which is considerably larger than reduction in transmission estimated for other respiratory pathogens. For example, Baker et al. (2020) estimated a 20% reduction in RSV transmission in the beginning of 2020. Future studies should explore whether the epidemiology and life history of Mp infections cause its transmission to be more susceptible to behavioral changes.

There are several limitations to our study. For example, we did not account for heterogeneity in Mp dynamics across states as the quantity of data did not allow for reliable model-fitting at a regional scale. We did not include age structure in the model as it would require more data. We also did not account for Mp strain dynamics, which have been hypothesized as another major driver of epidemic cycles (Kenri et al., 2008; Zhang et al., 2019). Finally, our estimate of NPI effects need to be interpreted with care, especially during a period with very little case data; the advantage of relying on a Bayesian framework is that these uncertainties can be captured and constrained using sensible priors. Despite these limitations, our prediction that a build up of a susceptible pool over the past 4 years will cause a large Mp outbreak is likely qualitatively robust.

So far, there have been limited modeling studies analyzing epidemic dynamics of Mp infections (Omori et al., 2015; Zhang et al., 2019). Our study reinforces recent work on the importance of understanding susceptible dynamics to predict the impact of perturbations to transmission (Baker et al., 2020; Park et al., 2024). As such, our analysis underlines the importance of serological surveillance data to capture the build up of the susceptible pool to foresee future outbreaks (Mina et al., 2020; Nguyen-Tran et al., 2022). Our study also underlines the potential of BIOFIRE@ Syndromic Trends network as a powerful surveillance measure.

## Data availability

All code is stored in a publicly available GitHub repository (https://github.com/parksw3/mycoplasma_pred).

## Supplementary Text

### Mathematical details

Here, we used the standard SIRS model to capture the dynamics of Mp epidemics. The dynamics of the model is governed by the following set of ordinary differential equations:

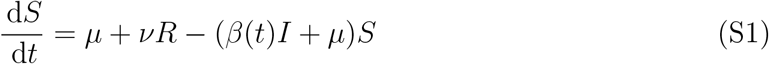

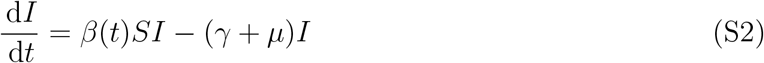

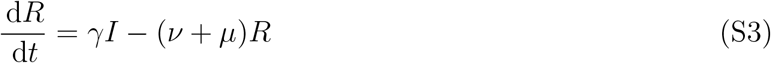

where *S, I*, and *R* represents the proportion of individuals in a susceptible, infected, and recovered compartment; *β*(*t*) represents time-varying transmission rate; *γ* represents the recovery rate; *ν* represents the immunity waning rate; and *μ* represents the birth and death rates, which are assumed to equal to keep the population size fixed. For simplicity, we assumed a population size of *S* + *I* + *R* = 1.

We discretized the model using the Euler scheme outlined in He et al. (2010) at a weekly time scale Δ*t* = 1 week:

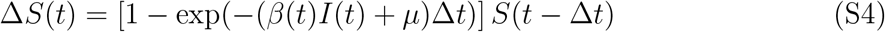

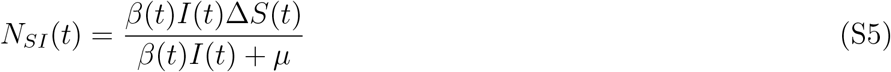

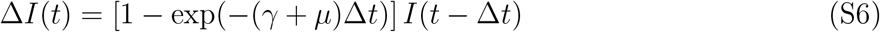

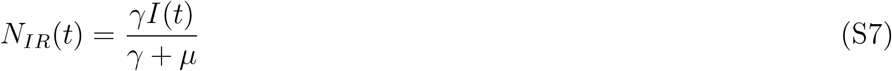

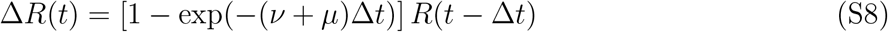

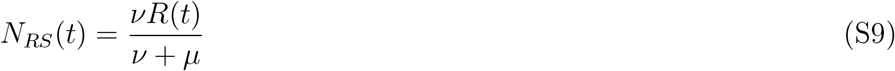

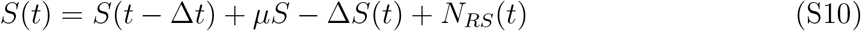

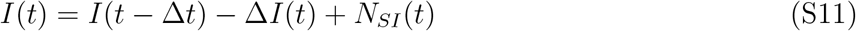

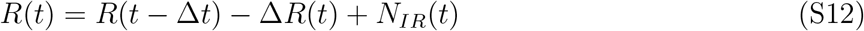

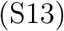

where Δ*X*(*t*) represents the total number of individuals leaving the compartment *X* at time *t*, and *N*_*XY*_ (*t*) represents the number of individuals leaving the compartment *X* to enter the compartment *Y* at time *t*. As explained in the main text, the transmission function was further decomposed into a periodic term *β*_seas_(*t*) and a non-periodic, time-varying term *δ*(*t*):

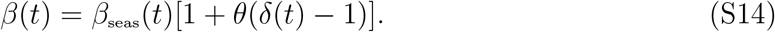

Here, the periodic term *β*_seas_(*t*) was modeled by estimating a weekly transmission rate for each of 52 weeks, which was modeled using cyclical, random-walk priors to allow for smoothing:

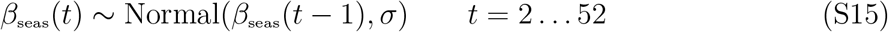

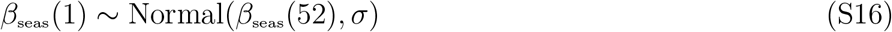

A half-normal prior was used for the standard deviation in smoothing *σ*:

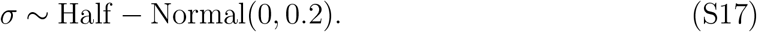

The non-periodic term was modeled by estimating a separate *δ* for every four weeks with a normal prior centered around 1:

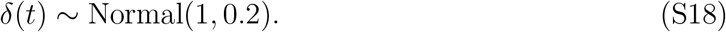

Finally, we assumed a lognormal error model to fit the model to observed positivity *P* (*t*):

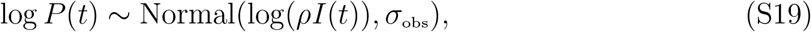

Here, *ρ* represents the scaling factor and *σ*_obs_ represents the residual standard deviation. For both terms, we used weakly informative half-normal priors to constrain the parameter space:

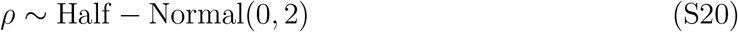

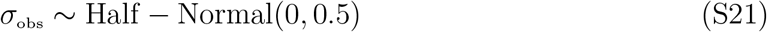

We also used weakly informative half-normal priors for the duration of immunity *τ* = 1*/ν*:

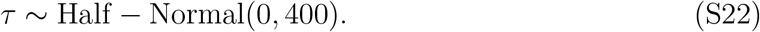

Finally, we assumed a uniform prior for the initial proportion susceptible *S*(0) and a half-normal prior for the initial proportion infected *I*(0):

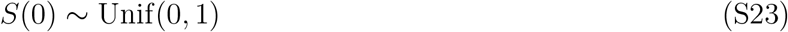

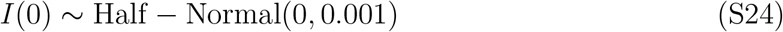

The model was fitted using rstan (Carpenter et al., 2017; Stan Development Team, 2024) with 4 chains, each containing 8000 iterations. Convergence was assessed based on the lack of warning signs from rstan, indicating: no divergent chains; no iterations exceeding maximum tree depth; sufficiently high Bayesian Fraction of Missing Information; sufficiently high effective samples sizes; and sufficiently low Rhat.

## Supplementary Figures

**Figure S1:**
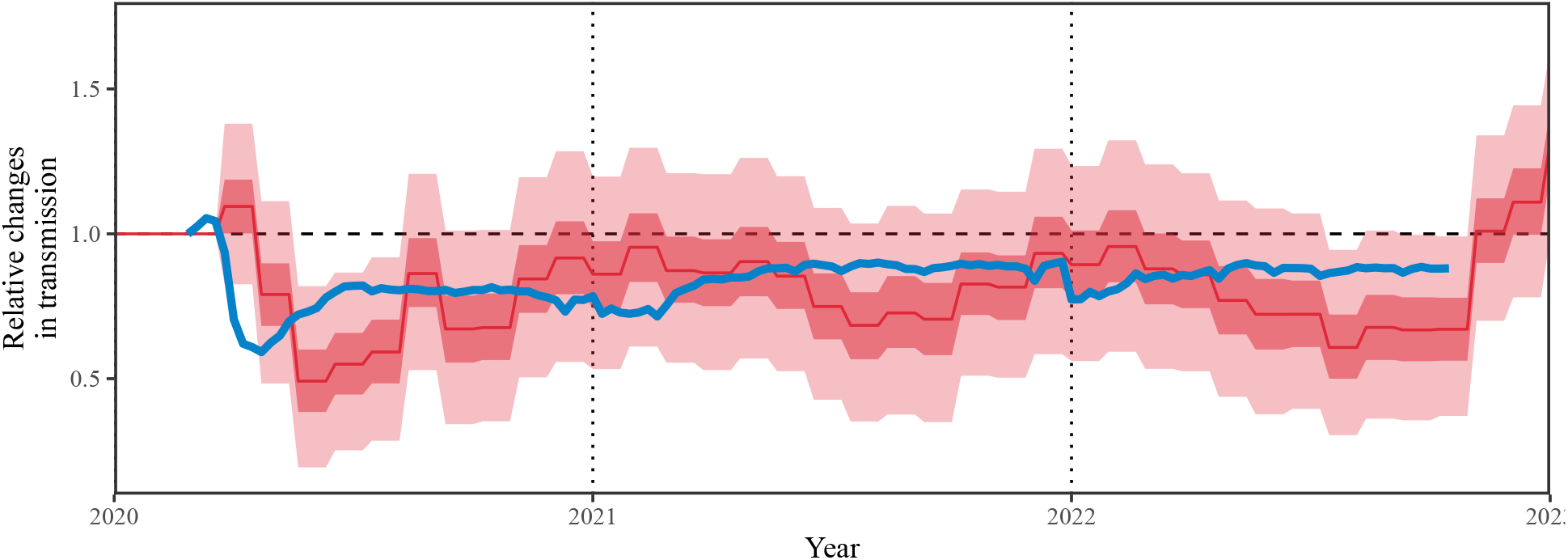
Comparisons of estimated changes in transmission (red) and mean Google mobility measures (blue). See Park et al. (2024) for the details of processing Google mobility data.

**Figure S2:**
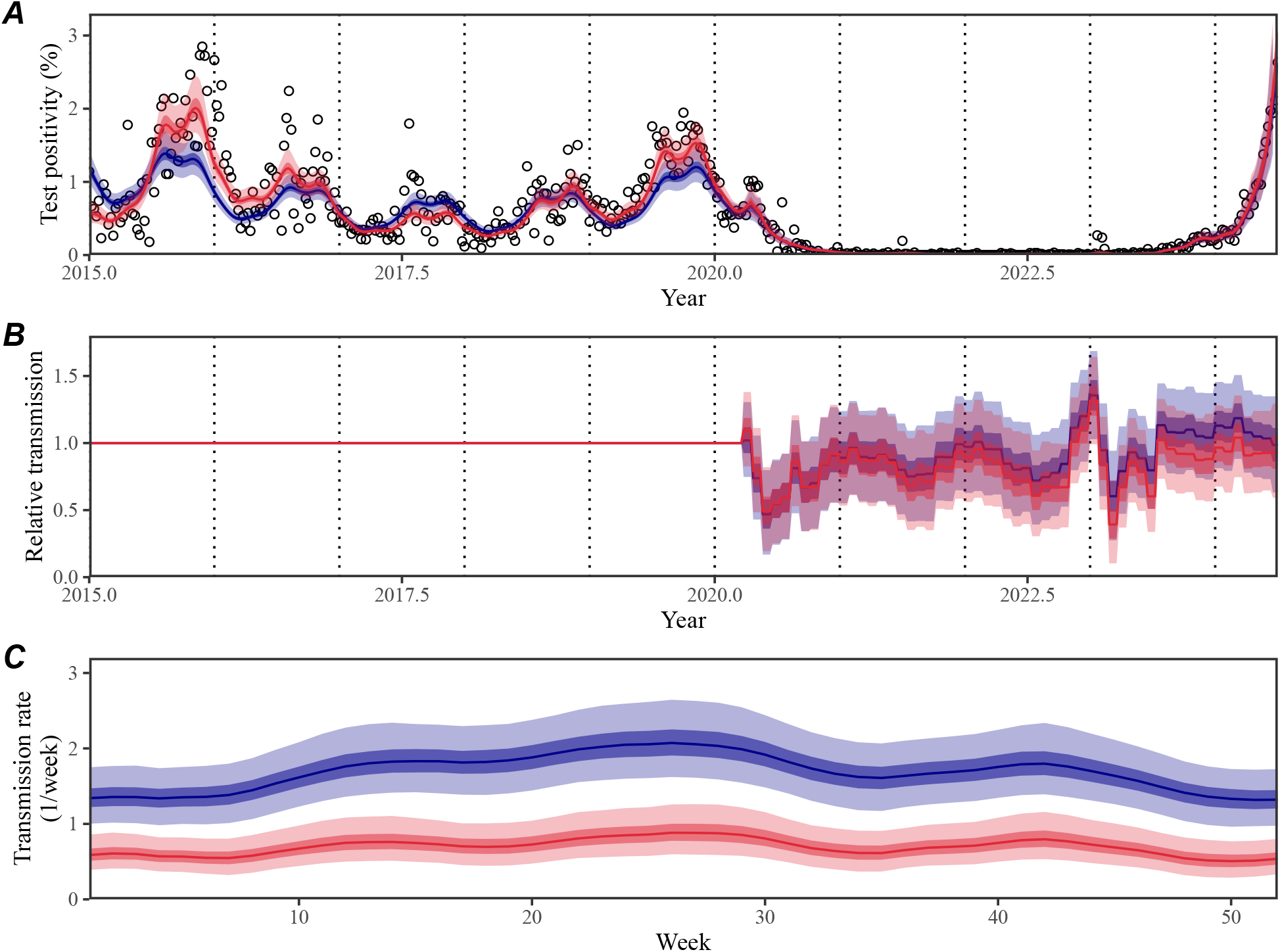
Comparisons of SIRS (red) and SIR (blue) model fits to Mycoplasma pneumoniae positivity in the US, 2015–2024. (A) Comparisons of observed (points) and fitted (line) changes in weekly test positivity for Mp infections. (B) Estimated non-periodic, time-varying transmission term, representing relative transmission *δ* following the introduction of NPIs. These changes are relative to the seasonal transmission rate shown in panel C; for example, 0.5 corresponds to a 50% reduction in transmission. (C) Estimated periodic transmission term *β*_seas_(*t*), representing seasonal transmission rate. Lines and shaded regions represent the estimated posterior median and corresponding 95% and 50% credible intervals.

